# COVID-19 vaccine equity and the right to health for displaced Venezuelans in Latin America

**DOI:** 10.1101/2022.10.21.22281363

**Authors:** David C. Hill, Zafiro Andrade-Romo, Karla Solari, Ellithia Adams, Lisa Forman, Daniel Grace, Alfonso Silva-Santisteban, Amaya Perez-Brumer

## Abstract

Given the magnitude of Venezuelan displacement in Latin America, there is a need to assess how migrants were, and will continue to be, addressed in COVID-19 vaccination policies. To explore migration status as a dimension of vaccine equity in Latin America and in relation to international human rights, we assessed national vaccination plans, peer-reviewed, and gray literature published between January 2020 and June 2021. Three key rights-related concerns were found to restrict the health rights of migrants in the region: 1) lack of prioritization of migrants in vaccine distribution; 2) onerous documentation requirements to be eligible for COVID-19 vaccination; and (3) how pervasive anti-migrant discrimination limited equitable health care access. While international human rights law prohibits against discrimination based on migration status, few countries analyzed realized their obligations to provide equal access to COVID-19 vaccines to non-citizens, including displaced Venezuelans. Especially for migrants and displaced people, effective and sustainable vaccination strategies for COVID-19 and future pandemics in Latin America must be guided not only by epidemiological risk but also seek to align with human rights obligations. To achieve this, States must also take special measures to facilitate vaccine access for communities facing systemic discrimination, exclusion, and marginalization.

## Introduction

In January 2021, Dr. Tedros Adhanom Ghebreyesus, Director-General of the World Health Organization (WHO), described the hoarding of COVID-19 vaccines by wealthy nations as a “catastrophic moral failure” that would “only prolong the pandemic, the restrictions needed to contain it, and human and economic suffering” [1]. Yet despite repeated calls for an equitable, globally coordinated approach to COVID-19 vaccination, vaccine apartheid has been unaddressed; as of March 2021, 78% of the 447 million doses were deployed in just ten countries [2]. By August 2021, over half of all people were fully vaccinated in Canada, the United States, the United Kingdom, and other Western countries, compared with just 11% in Latin America and the Caribbean [3–6]. These gross disparities are poised to deepen as third and fourth doses are increasingly recommended by governments and public health officials across North America and Europe [7]. Within countries already at the end of the line for doses, marginalized communities face additional barriers to access COVID-19 vaccines [8]. On both a global and a national scale, policymakers are grappling with how to prioritize COVID-19 vaccination to slow pandemic spread and protect the most vulnerable, including migrants.

Greater attention to migration status as a determinant of COVID-19 vaccine equity is urgently needed. COVID-19 vaccine access has repeatedly been denied to migrants, particularly those with irregular migration status, or those who entered a country without authorization or required documentation and therefore lack access to basic rights, including health care [9,10]. By May 2022, out of 180 countries evaluated globally by the United Nations International Organization for Migration, 39 countries excluded migrants with an irregular status from COVID-19 vaccination plans [11]. Yet, exclusion based on migration status violates international human rights prohibitions against “discrimination of any kind”, including nationality or migration status, raising important concerns regarding the health rights of migrants amid public health emergencies [12]. Indeed, in 2000 the Committee on Economic, Cultural, and Social Rights affirmed that “States are under the obligation to respect the right to health by, inter alia, refraining from denying or limiting equal access for all persons, including prisoners or detainees, minorities, asylum seekers and illegal immigrants, to preventive, curative and palliative health services” [13].

By December 2021, more than 6 million Venezuelans fled the country due to political, economic, and social turmoil, making it the second largest international displacement in contemporary history, behind only Syria’s refugee crisis [14]. Most Venezuelans remain in South America, placing severe socioeconomic strains on countries such as Argentina, Brazil, Chile, Colombia, Ecuador, and Peru, which collectively host over four million Venezuelans. While both regular and irregular migrants face stigmatization, discrimination, and health care access barriers, lack of documentation can make these issues particularly salient for irregular migrants. For many countries, government-issued identification documents (IDs) are required to access COVID-19 vaccinations. ID requirements coupled with heightened xenophobia and ongoing human rights violations may negatively impact how Venezuelan migrants are included in host country vaccination policies [15].

While the Latin American region has been significantly impacted by Venezuelan mass migration, limited research has assessed the sociopolitical climates shaping COVID-19 vaccine access for displaced Venezuelans. Thus, there is an urgent need to understand how rising numbers of displaced Venezuelans will be addressed in country-level COVID-19 vaccination policies. Addressing this gap, we examine the bioethical and human rights dimensions of country-level COVID-19 vaccination policies for migrants. We aim to better understand migration status (i.e. regular and irregular) as a dimension of vaccine equity through country-level vaccination plans and in relation to international human rights obligations and the “WHO SAGE values framework for the allocation and prioritization of COVID-19 vaccines”, which provides guidelines to promote equity within global and national prioritization of vaccination.

## COVID-19 vaccination facilities and equity frameworks

In recognition of underlining inequities pervasive in emergency vaccine administration, the “WHO SAGE values framework for the allocation and prioritization of COVID-19 vaccines” (hereafter referred to as the WHO/SAGE framework) was developed “for COVID-19 vaccines to contribute significantly to the equitable protection and promotion of human well-being among all people of the world” [16]. The WHO/SAGE framework is guided by a bioethical approach emphasizing that social vulnerabilities (including citizenship status) should be considered when prioritizing COVID-19 vaccination. However, as Sekalala and colleagues note, while the WHO/SAGE framework mentions human rights principles and references State obligations to promote ‘global equity’, it falls short of human rights obligations to hold States accountable to ensure equity in access to vaccines [17].

Second, the need for international collaboration led to the creation of COVID-19 Vaccines Global Access (COVAX) to support vaccine research and development and promote global equity in vaccine distribution [18]. COVAX contains a small “humanitarian buffer” (around 5 percent of doses), which includes doses for migrants and refugees who otherwise do not have access through national vaccination campaigns. Yet, the number of doses available through COVAX is wholly inadequate given the magnitude of the Venezuelan migration crisis and other migration crises [19]. COVAX does not specify how national prioritization of vaccination should proceed, leaving governments to define priority groups for COVID-19 vaccination.

Existing COVID-19 vaccination facilities and equity frameworks may not adequately account for the unique vulnerabilities facing migrants amid COVID-19. For example, the WHO/SAGE framework, which is meant to guide State actors, lacks accountability in upholding the health rights of non-citizens. Scholars have thus argued that COVID-19 vaccination frameworks can be strengthened by aligning with international human rights obligations that define health rights; this can create legally enforceable frameworks that can be used to hold States accountable for protecting rights, particularly amid emergencies such as COVID-19 [17]. Indeed, international human rights treaties (including the International Covenant on Civil and Political Rights and the International Covenant on Economic, Social, and Cultural Rights) are ratified and legally binding in most countries globally. The United Nations Committee on Economic, Social, and Cultural Rights has emphasized that access to COVID-19 vaccines is “an essential component of the right of everyone to the enjoyment of the highest attainable standard of physical and mental health and the right of everyone to enjoy the benefits of scientific progress and its applications” [20]. The Committee also underscores the obligation of States “as a matter of priority and to the maximum of their available resources, to guarantee all persons access to vaccines against COVID-19, without any discrimination” [20]. The integration of human rights into existing vaccination frameworks would transform ethical imperatives into obligations and rights, which can be mobilized in legal and political strategies by civil society and social actors to hold States accountable for ensuring health rights are fulfilled [17].

## Methods

Data are derived from a scoping review conducted between April and June 2021 evaluating the inclusion of Venezuelan migrants in COVID-19 vaccination policies in Latin America. Complete scoping review methods have been previously published [21]. The original scoping review broadly assessed COVID-19 vaccine access for displaced Venezuelans, while this secondary analysis explored the human rights dimensions of COVID-19 vaccination for displaced Venezuelans. For this analysis, country-level vaccination plans and statements from government officials were examined alongside international human rights obligations and the WHO/SAGE framework to explore how bioethical and human rights dimensions of COVID-19 vaccination were framed for Venezuelan migrants.

Four team members contributed to data extraction with at least one full text reviewer per data source. A search of peer-reviewed literature published between January 2020 and June 2021 yielded 142 results and 13 articles included in analysis; gray literature screening included 74 materials for full text review and 37 included in analysis, including publications and reports by nongovernmental organizations, multilateral agencies, and news media; and country-level documents were reviewed for 18 countries across Latin America and the Caribbean, with in-depth analysis for the six countries hosting most Venezuelans (Argentina, Brazil, Chile, Colombia, Ecuador, and Peru). Two team members reviewed Ministry of Health websites, policies, documents, and media related to COVID-19 vaccination plans.

Data were collected in English, Spanish, and Portuguese through a variety of online platforms for screening and data extraction (Covidence, Google Forms, Zotero, and Excel). Data extraction was divided by data source (peer-reviewed, country-level government policies, gray literature) with at least one full-text reviewer and data extractor per publication. Summative content analysis guided this analytic process to assess both what is explicit as well as implicit with regards to mechanisms for COVID-19 vaccine access based on migration status [22,23]. Research team coded sections of the documents into overarching interpretative themes and subthemes related to how migrants were explicitly described or implied in the sources and, in some cases, excluded from vaccination strategies. Codes were used to generate key themes across extracted documents and yielded summaries relevant to law, policy, and institutional practices for migrants. Emergent themes were discussed across the full team (6 members) who met twice weekly throughout the six-week study period to review extraction procedures and to compare, discuss, and interpret data.

## Results

Across the analysis, three key rights-related concerns were found to restrict the right to health for migrants in Latin America amid COVID-19: 1) lack of prioritization of migrants in vaccine distribution; 2) onerous documentation requirements to be eligible for COVID-19 vaccination; and 3) how pervasive anti-migrant discrimination limited equitable health care access. A detailed analysis of country-level vaccination plans, phases of vaccination, and ID requirements for vaccination is presented in Table 1, and Fig 1, in the Discussion, provides recommendations based on results for integrating human rights into existing COVID-19 vaccination facilities and frameworks (i.e. WHO SAGE/values).

**Table 1.**
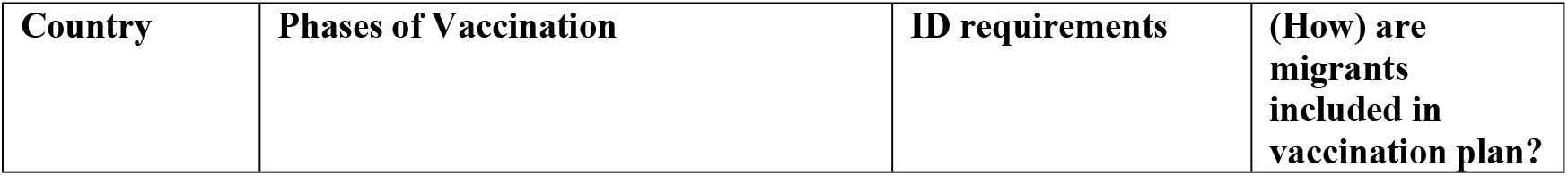
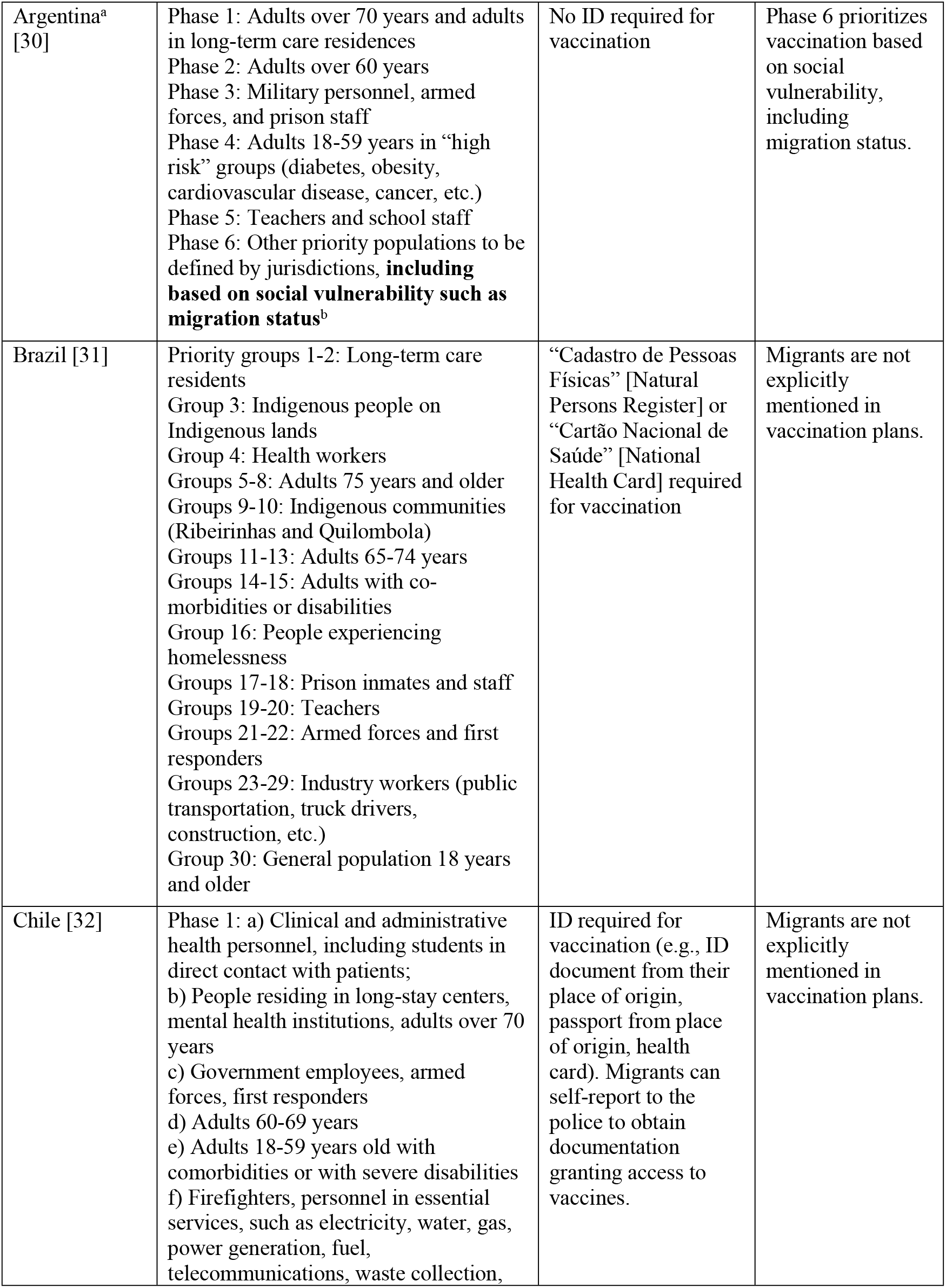
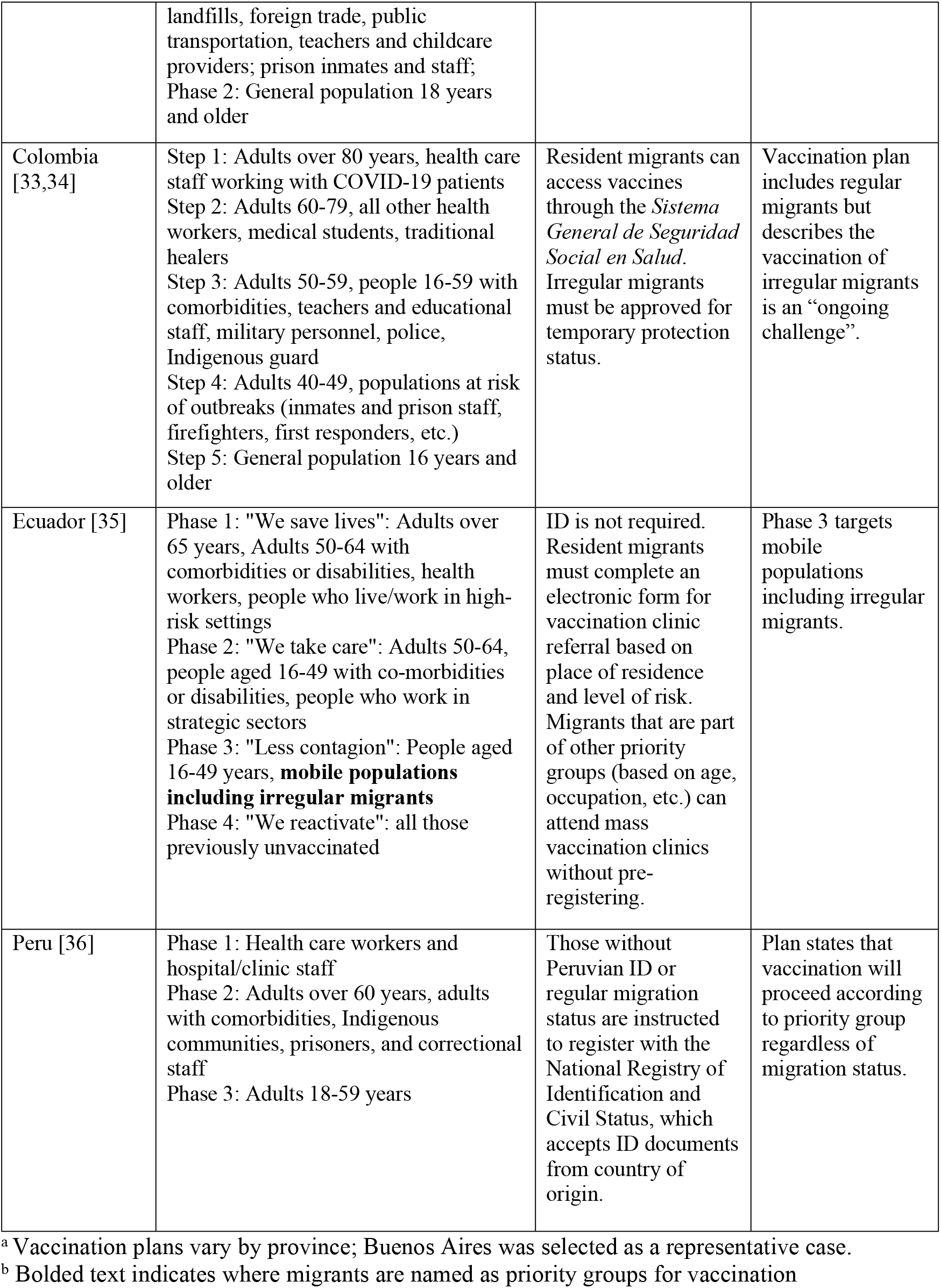
Phases of COVID-19 vaccination, ID requirements, and inclusion of Venezuelan migrants in six host countries as of June 2021

**Fig 1.**
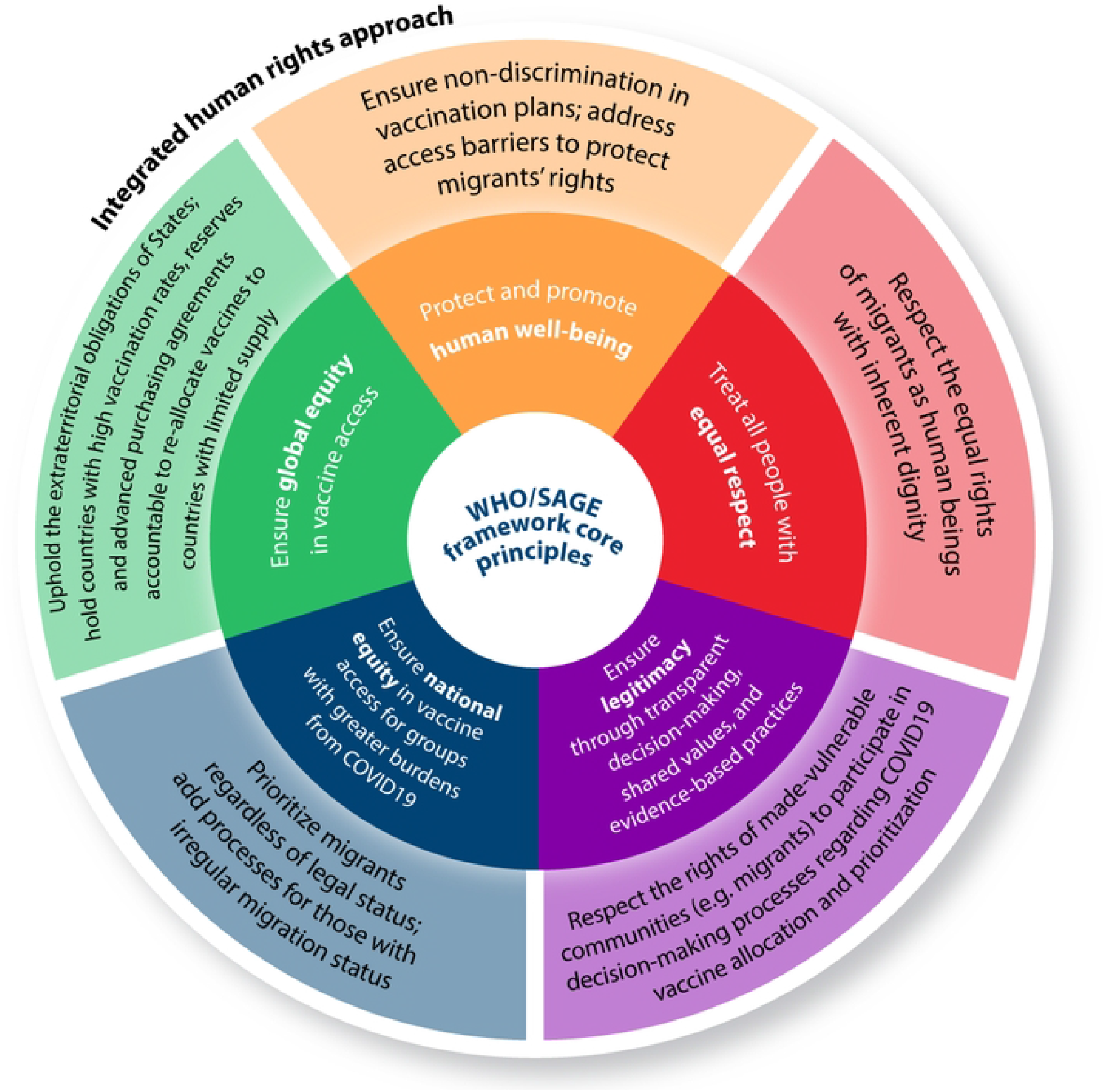
Integrating human rights into a modified WHO/SAGE framework to address the unmet COVID-19 needs of migrants (A) Summary of select principles in the WHO/SAGE framework. (B) Integrating human rights with WHO/SAGE to promote equitable access to COVID-19 vaccines for migrants.

### Lack of prioritization of migrants in vaccine distribution

Of the six countries reviewed in-depth, only Argentina and Ecuador identified migrants as priority groups for vaccination. In Argentina, migrants were prioritized after adults 60 and over, adults with comorbidities, military personnel, armed forces, prison staff, teachers, and school staff [24]. Similarly, in Ecuador, migrants were listed as a priority group for Phase 3 of the vaccination plan. While vaccination plans in both Argentina and Ecuador clarified that migrants may be eligible alongside citizens if they were part of other priority groups, these were defined by epidemiological risk factors with limited attention to structural factors such as overcrowded housing, precarious employment, and migration status. The first two phases of Ecuador’s vaccination plan, “we save lives” and “we take care”, prioritized adults 50 years and older, adults with comorbidities, and workers in strategic sectors [25]. Phase 3, which explicitly included migrants, was referred to as “limit contagion”, reinforcing stereotypes of migrants as disease vectors. Even when migrants were explicitly named as priority groups for vaccination, this was framed in terms of reducing risk for citizens.

Meanwhile, review of the gray literature revealed a rapidly shifting policy landscape that at times explicitly excluded migrants from vaccination efforts. In Chile, although the Constitution recognizes migrants’ right to health, the Minister of Foreign Affairs stated in February 2021 that irregular migrants would not be vaccinated against COVID-19 [26]. The decision was criticized by the Chilean Medical College, who argued that restricting COVID-19 vaccination based on migration status would disproportionately affect the most vulnerable [27]. In response, the Chilean government updated the policy to only restrict tourists from COVID-19 vaccination [28]. Similarly, in December 2020, Colombia sought to exclude irregular migrants from vaccination plans, including an estimated one million displaced Venezuelans with irregular migratory status [29]. The announcement triggered international outrage, leading President Duque to backtrack in February 2021 and announce a Temporary Protection Status (TPS) program that would allow migrants to formalize their status and thus access health care, including vaccination [29]. The discrimination evidenced in statements from government officials clearly juxtaposed human rights approaches that prohibit denying or limiting access to COVID-19 vaccines based on nationality or migration status.

While migrants may be eligible for vaccination based on age or occupation, country-level vaccination plans were often unclear if, how, and when non-citizens, particularly those without IDs, can access vaccines. Table 1 illustrates how reviewed countries allocated their limited COVID-19 vaccines to “at-risk” groups according to age, occupation, and co-morbidities, with limited information on how migrants and non-citizens can access COVID-19 vaccinations within these classifications.

In contrast with Argentina and Ecuador, which explicitly name migrants as priority groups, plans in Brazil, Chile, Colombia, and Peru were more ambiguous regarding access for migrants. Chile and Colombia defined priority groups predominantly by risk of morbidity and mortality (age and co-morbidities), as well as by occupation (health care and other “essential” workers). Yet, while migrants may be eligible based on age, occupation, or comorbidities, vaccination plans were often unclear regarding when and how migrants not eligible based on individual-level risk could access vaccines. Colombia’s plan indicated that while regular migrants could access vaccines alongside citizens, vaccination of irregular migrants was described as an “ongoing challenge” [33]. Some country-level approaches considered group vulnerability when, for instance, prioritizing vaccination in mental health institutions (Chile) and inmates and prison staff (Argentina, Brazil, Colombia, Chile, and Peru).

Findings from peer-reviewed and gray literature reveal how unstable housing, precarious employment, discrimination, and health care access barriers represent intersectional structural-level factors that constrain access to social protections for made-vulnerable communities. In Brazil and Peru, COVID-19 vaccination plans addressed structural factors that place Indigenous communities at increased vulnerability. For example, Brazil’s national plan notes collective ways of living coupled with long distances from health centers as factors that place Indigenous Quilombola and Ribeirinhas communities at increased vulnerability to COVID-19, and thus prioritizing these communities for COVID-19 vaccination [31]. Yet despite crowded living conditions, health care access barriers, language barriers, and precarious employment, which increase vulnerability to COVID-19 for many displaced Venezuelans in Brazil, migrants as a specific group are not prioritized within Brazil’s vaccination plan [37].

### Onerous documentation requirements to be eligible for COVID-19 vaccination?

While some vaccination plans included flexibility surrounding ID requirements for non-citizens or the acceptance of expired IDs, documentation requirements were noted as a recurrent vaccination barrier, particularly for migrants with irregular status [38]. As of March 2021, over 65 percent of Venezuelans in Colombia reported a lack of documentation as a health care access barrier [14]. In Colombia, while regular migrants can access vaccines with a residency permit, irregular migrants must apply for Temporary Protection Status (TPS). Yet, TPS applications are costly and can take months to process, further constraining or delaying COVID-19 vaccination [29]. Additional TPS eligibility criteria excluded migrants that arrived after January 31^st^, 2021 and those with a criminal record [14]. Zard et al. documented how relying on regularization processes as a route to vaccination delayed and constrained COVID-19 vaccine access for migrants and refugees [8]. To access vaccines in Colombia, Chile, and Brazil, displaced Venezuelans needed a passport, proof of legal status, or proof of enrolment in health care systems [31,33]. Registration for COVID-19 vaccination also often required booking an appointment online and uploading documentation, which the International Organization for Migration (IOM) has identified as a barrier for migrants who may lack computer access or internet connectivity [38]. Several support mechanisms were identified in the gray literature to reduce vaccination barriers, such as United Nations agencies in Brazil assisting with regularization applications, and temporary, transitory, or precarious residency permits being automatically extended in Argentina [39].

### How did pervasive anti-migrant discrimination limit equitable health care access?

Another crucial COVID-19 vaccination barrier evidenced across the peer-reviewed and gray literature was migrants’ distrust in health care systems, in part due to fear of deportation. The “UN joint guidance note on equitable access to COVID-19 vaccines for all migrants” calls on States to establish firewalls between health services and immigration authorities to prevent detention or deportation due to accessing health care services [40]. While an important call, limited accountability can further harm those most vulnerable. For example, in Chile, to access COVID-19 vaccines, displaced Venezuelans must self-report to police and disclose their irregular migration status [27]. Policies requiring disclosure of migration status can perpetuate the stigmatization of migrants, particularly as more than 100 migrants, mostly Venezuelans, were deported from Chile in February 2021. Then, in April 2021, another 55 migrants were deported as a part of the “Colchane plan” which sought to deport 1,800 migrants and increase border controls [10]. Similarly, in Peru, irregular migrants must register with the National Registry of Identification and Civil Status, and Brazil requests migrants register with the *Cadastro de Pessoas Físicas* (Naturalized Persons Registry) [36,41]. Amnesty International has called on States to “explicitly guarantee that vaccination will not be linked to legal status including ensuring that personal data gathered…for vaccination purposes will not be shared with law-enforcement agencies and used for immigration enforcement” [29]. Amid the fears and uncertainties following mass deportations, migrants may avoid registering and declaring their irregular migration status, thus limiting vaccine access.

Meanwhile, in 2021, amid increasing numbers of Venezuelans seeking to return to Venezuela in response to discrimination, xenophobia, and violence in host countries, Venezuelan President Maduro blamed returning migrants for upsurges in COVID-19 cases [42]. Indeed, the Office of the United Nations High Commissioner for Human Rights writes that “in situations of fear and uncertainty, such as the current pandemic, migrants can be particularly vulnerable to attitudes and behaviors that stigmatize and scapegoat them” [40]. Colombian President Duque justified the exclusion of Venezuelans from vaccination campaigns for fear that it would lead to a “stampede” of migrants rushing the border [43]. A senior government official in Venezuela even referred to migrants entering the country illegally as “biological weapons” [44]. Both across country-level policies and related gray literature, our review demonstrates that amid extreme resource constraints facing Latin American countries due to COVID-19 and Venezuelan mass migration, stigmatization and scapegoating can result in discriminatory policies that constrict migrants’ right to the highest attainable standard of health.

In addition to COVID-19 vaccine access specifically, results revealed health care access barriers more broadly for displaced Venezuelans. A report by the Mixed Migration Centre noted that 75 percent of displaced Venezuelans in Peru and 39 percent in Colombia believed they would not be able to access health care if they became ill with COVID-19 [45]. Similarly, compared with citizens, displaced Venezuelans in Chile were 7.5 times less likely to have access to health care [26]. Despite calls from the Pan American Health Organization for “health services [to] be inclusive and responsive to the needs of migrants” and to “[eliminate] geographical, economic, and cultural barriers”, our review revealed that displaced Venezuelans lacking formal documentation were disproportionately affected by health care access barriers [46]. While the integration of displaced Venezuelans into COVID-19 vaccination plans is an important step towards health equity, reviewed literature reinforce the need to address social and structural disparities that limit the right to health for migrants.

## Discussion

These findings advance understandings of migration status as a determinant of COVID-19 vaccine equity by presenting a regional analysis of how country-level COVID-19 vaccination policies in Latin America facilitate access for migrants and displaced people. While the scale and impact of Venezuelan mass migration were underscored in the peer-reviewed and gray literature reviewed, government plans and policies highlighted that States’ provided limited safeguards for non-citizens. Our results underscore the imperative to not only recognize but also address protections for non-citizens in Latin America. International human rights principles reinforce the obligations of States to ensure equity for all in access to COVID-19 vaccines, regardless of citizenship or migration status.

The potential impact of our results not only help underscore how existing gaps in COVID-19 vaccination could have been addressed by human rights approaches, but also advance understandings of how future public health interventions can better integrate human rights principles to complement strategies informed by epidemiological risk. Existing and pervasive disparities highlight how COVID-19 vaccines, no matter how effective at preventing infection or severe illness, cannot reach their full promise without the integration of context-specific sociopolitical factors in their rollout. Human rights-related concerns surrounding COVID-19 vaccine access for displaced Venezuelans include: lack of prioritization of migrants in vaccine distribution; onerous documentation requirements to be eligible for COVID-19 vaccination; and how pervasive anti-migrant discrimination limited equitable health care access. These findings align with reports from the United Nations International Organization for Migration and WHO that country-level vaccination plans often lack clarity regarding the inclusion of migrants and fail to address underlying health care access barriers, particularly for migrants with irregular status [47].

This research builds on existing literature urging the integration of human rights perspectives in responses to COVID-19 to ensure health equity for migrants [47]. We have identified key priorities when designing and implementing COVID-19 vaccination plans to ensure equity for migrants and/or displaced people in the Latin American region. Paralleling literature detailing the urgency of incorporating human rights perspectives into vaccine deployment, planning and coordination mechanisms must account for non-citizens, particularly those with irregular migration status [47]. More broadly, mechanisms need to be adopted to address pervasive discrimination and ensure secure and safe access to public health systems for migrants. Assessment of priority groups within country-level vaccination policies alongside gray and peer-reviewed literature allows for an examination of how governments ‘see’ migrants and can foster mechanisms to improve vaccine access. Fig 1 situates five of the principles guiding the allocation and prioritization of COVID-19 vaccination as described by the WHO/SAGE framework alongside recommendations for how existing gaps could have been addressed through the integration of human rights.

Recommendations based on this review build on the WHO/SAGE framework which outlines six principles guiding COVID-19 vaccine prioritization, and underscores how social vulnerabilities (including migration status) should inform national vaccination policies. These include: (1) human wellbeing; (2) equal respect; (3) global equity; (4) national equity; (5) reciprocity; and (6) legitimacy. The WHO/SAGE framework calls for migrants to be prioritized in COVID-19 vaccination plans given their increased social vulnerabilities and existing barriers in accessing health services. For example, the principle of “national equity” advocates for the prioritization of vulnerable communities, such as people living in poverty, refugees, and internally displaced persons, those affected by humanitarian emergencies, and irregular migrants. Recognizing their increased risk of COVID-19 acquisition and severe illness, the WHO/SAGE framework underscores why migrants *should* be named as priority groups for COVID-19 vaccination. Yet, as evidenced in our review, scant attention was paid to accountability in implementation of country-level vaccination plans, resulting in States prioritizing the vaccination of citizens over non-citizens.

Fig 1 builds on a modified WHO/SAGE Framework to present how COVID-19 vaccine allocation and prioritization can better consider the health rights of migrants and non-citizens. For example, while the existing WHO/SAGE framework suggests that all people should be treated with equal respect, human rights obligations hold States accountable for ensuring dignity and equal respect for all. Similarly, while the WHO calls for “a transparent consultation process” in decisions surrounding COVID-19 vaccine policies, a rights-based approach necessitates that made-vulnerable communities participate actively in decision-making processes. Finally, extraterritorial obligations underscore the responsibility of States to uphold the health rights of all people, not just those residing within their national borders. Reciprocity is a principle of narrower scope compared to the other five as it specifically addresses those within society “who bear substantial additional risks and burdens of COVID-19 response for the benefit of society” [16]. While many migrants are trained as health care workers and/or first responders, they are often unable to work in these fields given challenges with documentation, and thus do not qualify under this principle. Assessment of policies related to whether migrants and other displaced people can provide COVID-19 care irrelevant of documents was outside of the scope of our review, and thus reciprocity is not presented in Fig 1. Nonetheless, future research is needed to better understand how the principle of reciprocity can be interpreted and applied to migrants and other displaced people especially given known shortages of healthcare workers in the Latin American region [48].

While the WHO/SAGE framework underscores the need to consider social vulnerabilities, our review revealed that country-level vaccination plans rarely accounted for the structural inequities driving increased exposure to COVID-19 for marginalized communities. These findings support literature evidencing how unstable housing, precarious employment, discrimination, and health care access barriers represent intersectional structural-level factors that constrain access to social protections not only for migrants, but also for other marginalized groups [49]. Prioritizing vaccination based solely on epidemiological risk groups serves to individualize risk and invisibilize the structural-level factors that produce group vulnerabilities. As argued by Sekalala and colleagues, “international human rights law requires that ‘vulnerability’, if used as a criterion for priority access to COVID-19 vaccines, must include social vulnerability (e.g. socioeconomic status) in addition to medical vulnerability (e.g. comorbidities), and attend to intersectionalities” [17]. In this way, prioritization of COVID-19 vaccination must be informed not only by epidemiological risk of morbidity and mortality; a human rights perspective would also require social vulnerabilities be considered when defining vaccine priority groups. As shown by Freier et al., this position has been adopted by the Inter-American Commission on Human Rights and the Inter-American Court of Human Rights, who established that the human rights principles of equality and non-discrimination requires States to not only prohibit discrimination based on migration status, but also to take special measures to address and reverse structural and systemic discrimination, exclusion, and vulnerability faced by migrants [50]. In countries where migrants already faced high rates of discrimination, xenophobia, criminalization, poverty, exploitation, and violence, COVID-19 exacerbated inequities in employment, health, and access to social services [51].

Findings underscore that sustained attention must be paid to the structural inequities described above that are not new, but rather pervasive consequences of neoliberal globalization. This call to center vulnerability seeks to underscore the broader systemic oppressions that make certain communities in Latin America particularly vulnerable to COVID-19 and other epidemics. It demands attention to be paid to imperialist structural adjustment policies that have left public health systems underfunded and ill-equipped to deal simultaneously with mass migration and a pandemic. Indeed, in response to the emerging debt crisis in the 1980s, countries across Latin America accepted loans from the World Bank and International Monetary Fund; conditions of these loans included mandates to privatize state assets, cut government spending, devalue currencies, and liberalize trade [52]. These large-scale cuts to government spending on social services widened health inequities as health care became increasingly unaffordable for the most vulnerable, including migrants [52]. The prioritization of COVID-19 vaccines in Latin America must be situated within ongoing neoliberal reforms yielding chronically underfunded social systems, including health care [53]. In sum, COVID-19 has repeatedly revealed the porousness of borders, yet responses continue to neglect the globalized and interconnected nature of the pandemic.

Caution should be observed in the interpretation of these results as findings are inherently limited by dynamic changes in approaches to COVID-19 vaccination that have occurred since this scoping review was conducted between April and June 2021. Indeed, review of country-level vaccination plans revealed at times daily shifts in vaccine prioritization and allocation, as well as migrants’ eligibility for vaccination, underscoring how rights claims were contested, mediated, and resolved across the study period. That said, given the scope of our three-month data collection period, it was not possible to fully assess how COVID-19 vaccination policies shifted across time. Additionally, vaccination data published across Ministry of Health websites did not disaggregate based on migration status, so the effects of COVID-19 vaccination policies on access to vaccines in practice could not be assessed. Further attention is also needed to the phases of vaccination, as well as definition of priority groups across different settings, not only in initial doses, but in the administration of booster doses also. Nonetheless, this review underscores ongoing gaps in COVID-19 vaccination plans as well as the crucial need to uphold international human rights obligations and consider the unique social vulnerabilities of displaced Venezuelans. As Peru and Brazil have done for Indigenous communities, this could be accomplished through an approach to vaccination that considers not just individual-level risk factors but also intersectional, structural-level factors that place displaced Venezuelans at increased risk for COVID-19. Ensuring equitable access to COVID-19 vaccines is thus a first - and an essential - step to respond to COVID-19 more equitably and prepare for future public health crises in the context of displaced populations.

## Conclusion

Amid rapidly evolving COVID-19 vaccination plans, findings presented here highlight how anti-migrant discrimination can be reinforced by governments and health systems when the human rights of migrants are not accounted for in vaccination policies across Latin America. Countries across the region prioritized the allocation of limited vaccines to ‘at-risk’ groups as defined by countries and guided by primarily epidemiological factors (age, occupation, co-morbidities, etc.). Limited attention to social vulnerabilities (such as housing instability, economic precarity), and global forces driving the migration crisis (such as imperialism and neoliberalism), has exacerbated deep-rooted structural inequities faced by those abandoned by social safety nets, such as migrants. Keen attention and deeper inquiry are needed to the unique barriers faced by Venezuelan migrants with irregular status, defined as those who entered a country without the necessary authorization or documentation. Requirements for identification and other government-issued documentation to access vaccines creates critical access barriers (e.g. fear of deportation and xenophobic harassment, lost/expired documents), particularly for migrants with irregular status. Meanwhile, issues of cost, lack of familiarity with health care systems, and stigma and discrimination are intersectional issues that may be more salient among irregular migrants, particularly amid COVID-19 and future pandemics [54]. Stigmatization and scapegoating can result in discriminatory policies that further limit the ability of migrants to achieve the “highest attainable standard of health”. The integration of human rights, rooted in legally binding treaties that can be used in litigation and advocacy, into public health responses is a key strategy to improve accountability within implementation and uphold the health rights of marginalized and made-vulnerable communities. To ensure health equity for migrants and displaced people, future COVID-19 vaccination strategies in Latin America must be informed not just by epidemiological approaches, but also by international human rights.

## Data Availability

The data that support the findings of this study are available from the corresponding author, DCH, upon reasonable request.

## Acknowledgements

This work was supported by the Canadian Social Sciences and Humanities Research Council (SSHRC), Insight Development Grant.

## Notes

### Competing Interest Statement

The authors have declared no competing interest.

### Author Declarations

IRB not required. This project was a scoping review of the literature.

## References

1. World Health Organization. WHO Director-General’s opening remarks at 148th session of the Executive Board [Internet]. 2021. Available from: https://www.who.int/director-general/speeches/detail/who-director-general-s-opening-remarks-at-148th-session-of-the-executive-board

2. Office of the United Nations High Commissioner for Human Rights. UN expert says global coordination and more equitable sharing of COVID-19 vaccines key to recovery [Internet]. 2021 [cited 2021 Aug 26]. Available from: https://www.ohchr.org/EN/NewsEvents/Pages/DisplayNews.aspx?NewsID=26683&LangID=E

3. Government of the United Kingdom. Vaccinations in the UK | Coronavirus in the UK [Internet]. 2021 [cited 2021 Aug 27]. Available from: https://coronavirus.data.gov.uk/details/vaccinations

4. CDC. COVID Data Tracker [Internet]. Centers for Disease Control and Prevention. 2020 [cited 2021 Aug 27]. Available from: https://covid.cdc.gov/covid-data-tracker

5. Government of Canada. Vaccines for COVID-19 [Internet]. 2020 [cited 2021 Aug 27]. Available from: https://www.canada.ca/en/public-health/services/diseases/coronavirus-disease-covid-19/vaccines.html

6. The Lancet. COVID-19 in Latin America—emergency and opportunity. The Lancet. 2021 Jul;398(10295):93.

7. John T. The definition of “fully vaccinated” is changing to three covid-19 doses [Internet]. CNN. 2021 [cited 2021 Dec 6]. Available from: https://www.cnn.com/2021/11/17/world/coronavirus-newsletter-intl-17-11-21/index.html

8. Zard M, Lau LS, Bowser DM, Fouad FM, Lucumí DI, Samari G, et al. Leave no one behind: ensuring access to COVID-19 vaccines for refugee and displaced populations. Nat Med [Internet]. 2021 Apr 19 [cited 2021 May 13]; Available from: http://www.nature.com/articles/s41591-021-01328-3

9. International Organization for Migration. Glossary on Migration [Internet]. 2011 [cited 2021 Dec 6]. Available from: https://publications.iom.int/system/files/pdf/iml25_1.pdf

10. R4V. Situation report of the activities implemented in January and February 2021 by the members of the R4V Venezuela situation coordination platform in Chile. [Internet]. 2021. Available from: https://data2.unhcr.org/es/documents/details/85832

11. International Organization for Migration. Migration inclusion in COVID-19 vaccination deployment [Internet]. 2022 Mar [cited 2022 Oct 10]. Available from: https://www.iom.int/sites/g/files/tmzbdl486/files/our_work/DMM/Migration-Health/IOM-Vaccine-Inclusion-Mapping-17-May-2021-global.pdf

12. International Covenant on Economic, Social, and Cultural Rights (ICESCR). G.A. Res. 2200A (XXI). 1966.

13. United Nations Committee on Economic, Cultural, and Social Rights. General Comment No. 14: The right to the highest attainable standard of health (Art.12). 2000.

14. Quarterly Mixed Migration Update [Internet]. Mixed Migration Centre. 2021 [cited 2021 May 28]. Available from: https://mixedmigration.org/resource/quarterly-mixed-migration-update-lac-q1-2021/

15. Human Rights Watch. Brazil: Events of 2020. In: World Report 2021 [Internet]. 2020 [cited 2021 Nov 17]. Available from: https://www.hrw.org/world-report/2021/country-chapters/brazil

16. World Health Organization. WHO SAGE values framework for the allocation and prioritization of COVID-19 vaccination [Internet]. 2020 Sep [cited 2021 Sep 16]. Available from: https://apps-who-int.myaccess.library.utoronto.ca/iris/bitstream/handle/10665/334299/WHO-2019-nCoV-SAGE_Framework-Allocation_and_prioritization-2020.1-eng.pdf?ua=1

17. Sekalala S, Perehudoff K, Parker M, Forman L, Rawson B, Smith M. An intersectional human rights approach to prioritising access to COVID-19 vaccines. BMJ Global Health. 2021 Feb 1;6(2):e004462.

18. World Health Organization. COVAX: Working for global equitable access to COVID-19 vaccines. [Internet]. [cited 2021 Aug 31]. Available from: https://www.who.int/initiatives/act-accelerator/covax

19. Berkley S. COVAX explained [Internet]. GAVI The Vaccine Alliance. 2020 [cited 2021 May 25]. Available from: https://www.gavi.org/vaccineswork/covax-explained

20. United Nations Committee on Economic, Social, and Cultural Rights. Statement on universal affordable vaccination against coronavirus disease (COVID-19), international cooperation and intellectual property. [Internet]. 2021. Available from: https://digitallibrary.un.org/record/3921880?ln=en#record-files-collapse-header

21. Perez-Brumer A, Hill D, Andrade-Romo Z, Solari K, Adams E, Logie C, et al. Vaccines for all? A rapid scoping review of COVID-19 vaccine access for Venezuelan migrants in Latin America. Journal of Migration and Health. 2021 Jan 1;4:100072.

22. Hsieh HF, Shannon SE. Three Approaches to Qualitative Content Analysis. Qual Health Res. 2005 Nov;15(9):1277–88.

23. Schaaf M, Boydell V, Topp SM, Iyer A, Sen G, Askew I. A summative content analysis of how programmes to improve the right to sexual and reproductive health address power. BMJ Glob Health. 2022 Apr;7(4):e008438.

24. Ministerio de Salud. Resolución 2883/2020 [Internet]. Buenos Aires, Argentina; 2020. Available from: https://www.argentina.gob.ar/normativa/nacional/disposici%C3%B3n-1904-2020-336399/texto

25. Ministerio de Salud Pública. Plan de vacunacion 9 100, Respuestas a inquietudes ciudadanas sobre el plan de vacunacion 9/100 [Internet]. 2021. Available from: https://www.salud.gob.ec/wp-content/uploads/2021/06/31-05-2021_-Preguntas-y-Respuestas_Plan-de-Vacunacion-9100_validado.pdf

26. Cabieses B, Oyarte M. Acceso a salud en inmigrantes: identificando brechas para la protección social en salud. Rev Saude Publica. 54(20):1–13.

27. Andina AP de N. Covid-19: Chile da marcha atrás y aclara que vacunación incluye a extranjeros irregulares [Internet]. 2021 [cited 2021 Jun 2]. Available from: https://andina.pe/agencia/noticia-covid19-chile-da-marcha-atras-y-aclara-vacunacion-incluye-a-extranjeros-irregulares-833441.aspx

28. Vargas F. Minsal recalca que solo extranjeros que realizan turismo no pueden vacunarse contra el covid-19 | Emol.com [Internet]. Emol. 2021 [cited 2021 Jun 2]. Available from: https://www.emol.com/noticias/Nacional/2021/05/31/1022465/Minsal-Vacunacion-Extranjeros.html

29. Rahhal N. Refugees and COVID-19 Vaccinations: Part of the Solution but Not Always Part of the Plan. Amnesty International. 2021 May 12;4.

30. Gobierno de la Provincia de Buenos Aires. Plan provincial público, gratuito y optativo contra COVID-19 [Internet]. Vacunarnos es cuidarnos entre todos. 2021. Available from: https://vacunatepba.gba.gob.ar/

31. Ministério da Saúde, Secretaria de Vigilância em Saúde. NOTA TÉCNICA N° 155/2021-CGPNI/DEIDT/SVS/M [Internet]. Brazil: Ministério da Saúde; N/A [cited 2021 Oct 23] p. 6. Available from: https://www.gov.br/saude/pt-br/media/pdf/2021/marco/16/nt_155-2021-cgpni_priorizacao_grupos.pdf

32. Gobierno de Chile. Grupos objetivos para la vacunación contra SARS-CoV-2 según el suministro de vacunas [Internet]. 2021. Available from: https://www.minsal.cl/wp-content/uploads/2021/03/GRUPOS-OBJETIVOS-3-marzo-2021.pdf

33. Ministerio de Salud y Protección Social. Plan Nacional de Vacunación contra el Covid-19 [Internet]. Bogotá; 2021 Feb. Report No.: 2. Available from: https://mivacuna.sispro.gov.co/MiVacuna?v1

34. Ministerio de Relaciones Exteriores. Decreto 216 Por medio del cual se adopta el Estatuto Temporal de Protección para Migrantes Venezolanos [Internet]. Bogotá D.C., Colombia; 2021. Available from: https://www.migracioncolombia.gov.co/normas/por-medio-del-cual-se-adopta-el-estatuto-temporal-de-proteccion-para-migrantes-venezolanos-decreto-216-del-1-de-marzo-de-2021

35. Jumbo B. Acnur y el gobierno de Ecuador definen el mecanismo para vacunar a los extranjeros en condición de movilidad como venezolanos y colombianos [Internet]. El Comercio. 2021 [cited 2021 Jun 17]. Available from: https://www.elcomercio.com/actualidad/ecuador/vacunacion-extranjeros-venezolanos-covid-ecuador.html

36. Gobierno de Perú. Resolución ministerial no 488: Plan nacional actualizado de vacunación contra la covid-19 [Internet]. 2020. Available from: https://cdn.www.gob.pe/uploads/document/file/1805113/Plan%20Nacional%20Actualizado%20contra%20la%20COVID-19.pdf

37. Integration of Venezuelan Refugees and Migrants in Brazil - Brazil [Internet]. ReliefWeb. 2021 [cited 2021 May 25]. Available from: https://reliefweb.int/report/brazil/integration-venezuelan-refugees-and-migrants-brazil

38. International Organization for Migration. Migrant inclusion in COVID-19 vaccination Campaigns [Internet]. 2021 May [cited 2021 Jun 11] p. 1–20. Available from: https://www.iom.int/sites/default/files/our_work/DMM/Migration-Health/iom-vaccine-inclusion-mapping-17-may-2021-global.pdf

39. Peduzzi P. Operación Acogida ya reubicó a 50.000 refugiados venezolanos [Internet]. Agência Brasil. 2021 [cited 2021 Jun 7]. Available from: https://agenciabrasil.ebc.com.br/es/direitos-humanos/noticia/2021-04/operacion-acogida-ya-reubico-50000-refugiados-venezolanos

40. Office of the United Nations High Commissioner for Human Rights. Joint guidance note on Equitable Access to COVID-19 Vaccinations for All Migrants. 2021. Available from: https://www.ohchr.org/Documents/Issues/Migration/JointGuidanceNoteCOVID-19-Vaccines-for-Migrants.pdf

41. Ministério da Saúde, Governo do Brazil. Perguntas e respostas [Internet]. 2021 [cited 2022 Oct 4]. Available from: https://www.gov.br/saude/pt-br/coronavirus/perguntas-e-respostas

42. Standley CJ, Chu E, Kathawala E, Ventura D, Sorrell EM. Data and cooperation required for Venezuela’s refugee crisis during COVID-19. Globalization and Health. 2020 Oct 22;16(1):103.

43. Parkin Daniels J. Alarm at Colombia plan to exclude migrants from coronavirus vaccine [Internet]. the Guardian. 2020 [cited 2021 Jun 28]. Available from: http://www.theguardian.com/global-development/2020/dec/22/colombia-coronavirus-vaccine-migrants-venezuela-ivan-duque

44. Washington Post. As coronavirus explodes in Venezuela, Maduro’s government blames ‘biological weapon’: the country’s returning refugees. [Internet]. [cited 2021 Nov 9]; Available from: https://www.washingtonpost.com/world/the_americas/coronavirus-venezuela-migrant-maduro/2020/07/19/582c659c-c518-11ea-a99f-3bbdffb1af38_story.html

45. Mixed Migration Centre. Access to health services for Venezuelans in Colombia and Peru during the COVID-19 pandemic [Internet]. 2021 [cited 2021 May 28]. Available from: https://mixedmigration.org/resource/4mi-snapshot-access-to-health-services-for-venezuelans-in-colombia-and-peru-during-the-covid-19-pandemic/

46. Pan American Health Organization. Resolution CD55/11, Rev 1. Health of migrants [Internet]. [cited 2021 May 24]. Available from: https://www.paho.org/en/documents/cd5511-rev-1-health-migrants

47. World Health Organization. COVID-19 immunization in refugees and migrants: principles and key consideration [Internet]. 2021 Aug [cited 2022 Jul 17]. Available from: https://www.who.int/publications/i/item/covid-19-immunization-in-refugees-and-migrants-principles-and-key-considerations-interim-guidance-31-august-2021

48. Institute for Health Metrics and Evaluation. Worldwide shortage of health workers threatens effective health coverage [Internet]. [cited 2022 Aug 15]. Available from: https://www.prnewswire.com/news-releases/worldwide-shortage-of-health-workers-threatens-effective-health-coverage-301553411.html

49. Homan P, Brown TH, King B. Structural intersectionality as a new direction for health disparities research. Journal of Health and Social Behavior. 2021 Sep 1;62(3):350–70.

50. Freier LF, Aron Said V, Quesada Nicoli D. Non-discrimination and special protection for migrants and refugees. International Journal of Discrimination and the Law. 2022 Aug 5;13582291221115538.

51. Office of the United Nations High Commissioner for Human Rights. A pandemic of exclusion: The impact of COVID-19 on the human rights of migrants in Libya [Internet]. 2021. Available from: https://www.ohchr.org/Documents/Issues/Migration/A_pandemic_of_exclusion.pdf

52. Birn AE, Nervi L, Siqueira E. Neoliberalism redux: The global health policy agenda and the politics of cooptation in Latin America and beyond: Debate: The cooptation of global health in Latin America. Development and Change. 2016 Jul;47(4):734–59.

53. Vasquez EE, Perez-Brumer A, Parker RG, editors. Social inequities and contemporary struggles for collective health in Latin America. Global Public Health. 2019;14(6–7).

54. Maestripieri L. The COVID-19 pandemics: why intersectionality matters. Frontiers in Sociology [Internet]. 2021 [cited 2022 Mar 10];6. Available from: https://www.frontiersin.org/article/10.3389/fsoc.2021.642662

